# Association of Epigenetic Age Acceleration with Early Stage Heart Failure: Sex-Specific Effects in the Project Baseline Health Study

**DOI:** 10.1101/2024.12.18.24319187

**Authors:** Perisa Ashar, Nicholas Cauwenberghs, Kalyani Kottilil, Ranee Chatterjee, Neha Pagidipati, Pamela S. Douglas, R. Scooter Plowman, Adrian F. Hernandez, Kenneth W. Mahaffey, Francois Haddad, Svati H. Shah, Lydia Coulter Kwee, the Project Baseline Health Study Group

## Abstract

**Background:** Epigenetic age acceleration (EAA), defined as blood DNA methylation-derived biological age exceeding chronological age, has emerged as a potential biomarker of cardiovascular health and disease. EAA has been associated with symptomatic heart failure (HF); however, its connection to early-stage HF remains unclear. Moreover, the association of EAA with cardiovascular disease may differ by sex.

**Methods:** We analyzed participants from the Project Baseline Health Study, a deeply-phenotyped longitudinal cohort. HF staging was performed using echocardiographic measures and clinical criteria, classifying participants into stage 0/B0 (no HF risk factors), stage A (HF risk factors only, without structural cardiac abnormalities), and stage B1 (structural changes without HF symptoms). EAA was calculated as the residual of Horvath methylation epigenetic age regressed on chronological age. We compared EAA across HF stages using ANOVA models adjusted for sex, and in secondary analyses, investigated associations between EAA and echocardiographic parameters as well as sex-specific differences.

**Results:** Among 1,338 participants (mean age: 49.2±15.4 years; 53.5% female), 41.7% were stage 0/B0 (n=558), 39.4% were stage A (n=527), and 18.9% were stage B1 (n=253). EAA tended to differ across HF stages (p=0.07), with higher EAA in stage B1 participants compared to those in stage 0/B0 or A. Sex-stratified models suggested that this trend was more pronounced in males than females. Among individual echocardiographic parameters, greater left ventricular posterior wall thickness was associated with higher EAA (p=0.046). In analyses restricted to stage A and B1 participants, higher EAA in B1 approached significance only in males (p=0.051).

**Conclusions:** These findings suggest that elevated EAA may occur even in the early, asymptomatic stages of HF, potentially reflecting underlying biological aging processes associated with early cardiac structural abnormalities. The effects may be sex-specific, with males showing a stronger relationship between EAA and stage B1 HF. Although validation through larger, longitudinal studies is necessary, our results support EAA as a potential biomarker for early cardiovascular risk assessment and underscore the need of considering sex differences in early HF pathophysiology.

## Introduction

Epigenetic age acceleration (EAA) occurs when biological age, as measured by blood DNA methylation, is greater than chronological age, and has emerged as a potential biomarker for cardiovascular health. Prior studies have shown that positive EAA is associated with increased burden of cardiovascular risk factors, cardiovascular disease including heart failure, and allcause mortality (1-3), but its relationship with early-stage heart failure remains unexplored. Further, recent evidence suggests that EAA may exhibit sex-specific patterns in cardiovascular disease, though the mechanisms underlying these differences remain unclear (4). Thus, we investigated the association between EAA with early stage (or stage B) heart failure (HF) in the Project Baseline Health Study (PBHS) cohort, with a focus on potential sex-specific differences.

## Methods

### Study Design and Heart Failure Staging

The PBHS study design has been reported (5). Briefly, PBHS enrolled 2502 participants from four sites and obtained medical histories, performed deep clinical assessments and collected biospecimens annually. Cardiac echocardiograms were performed at baseline and read by board-certified echocardiographers in a core lab, and in combination with self-reported symptoms and medical diagnoses, were used to define stages of HF (6). Participants without HF risk factors, cardiac structural abnormalities (e.g. left ventricular hypertrophy) or HF symptoms were designated as stage 0; participants were classified as stage A if they had HF risk factors, but no cardiac structural abnormalities or HF symptoms. Stage B HF was defined as the presence of cardiac structural abnormalities in the absence of symptoms of HF and was further separated into B0 and B1 subgroups (without and with accompanying HF risk factors, respectively); for this analysis, B0 was grouped with stage 0. Stage C HF was defined as prior or current symptoms of HF with evidence of cardiac structural abnormalities (7). Participants with stage C (overt HF symptoms) were excluded.

### DNA Methylation Profiling and Epigenetic Age Calculation

Methylation profiling using Illumina EPIC 850k arrays was performed on available enrollment samples in the Verily core labs using standard methods and as described previously (3). Briefly, DNA was extracted from frozen whole blood and, after quantification, underwent automated bisulfite conversion, desulphonation, bead-based cleanup, and elution using an EZ-96 DNA MethylationLightning MagPrep kit (Zymo Research, Irvine, CA, USA) per the manufacturer’s protocol. The bisulfite-converted DNA was processed through standard Illumina Infinium MethylationEPIC microarray assay protocols (Illumina Inc., San Diego, CA, USA), and the resulting raw data were processed using *minfi* (8) to produce methylation estimates, QC metrics, and detection p-values. The Horvath epigenetic age was calculated (9), which combines methylation levels at 353 sites across the genome. EAA was calculated as the residual between chronological age and the calculated Horvath methylation epigenetic age, as described previously (3).

### Statistical Analysis

ANOVA models were used to test for global associations between HF stages (0/B0, A and B1) and EAA, first adjusting for sex, and then in sex-stratified models. In secondary analyses, we explored associations between EAA and the cardiac structural components of stage B HF using two approaches: 1) linear models regressing EAA on individual structural components of stage B HF and 2) models restricted to stage A and stage B1 participants.

### Data Availability and Ethical Approval

Deidentified PBHS data corresponding to this study are available upon request for the purpose of examining its reproducibility, with requests subject to approval by PBHS governance. All participants were appropriately consented for use of biospecimens and data. Informed consent was obtained from all participants enrolled in PBHS in accordance with the Helsinki declaration. The study was approved by a central Institutional Review Board (the WCG IRB; approval tracking number 20170163, work order number 1-1506365-1) and IRBs at each of the participating institutions (Stanford University, Duke University, and the California Health and Longevity Institute).

## Results

Of the 2502 participants enrolled in PBHS, 2071 had HF staging available at enrollment, while 1661 had methylation profiling. After removing 35 participants with stage C HF, 1338 participants with both HF phenotyping and methylation profiling were included in analysis: 558 (41.7%) with stage 0/B0 HF, 527 (39.4%) with stage A, and 253 (18.9%) participants with stage B1 (**Table 1**). The mean chronologic age increased progressively across stages (stage 0/B0: 41.2±14.0 years; stage A: 51.4±15.0 years; stage B1: 58.1±14.2 years), with balanced sex distribution (females: 53.0%, 53.9%, and 54.2% respectively). There was a higher percentage of individuals of self-reported Black race in stage B1 as compared with stage A or 0/B0 HF (30.0% [B1] vs. 23.5% [A] and 9.7% [0/B0]), whereas self-reported Asian individuals had a higher percentage of stage 0/B0 as compared with stage A or B1 HF (18.3%, 6.8%, and 7.5%, respectively). As expected, there was a higher burden of general cardiovascular risk factors in stage B1 HF (mean body mass index [BMI] 32.25±6.81 [stage B1] kg/m^2^ vs. 31.11±6.58 kg/m^2^ [stage A] vs. 24.23±2.88 kg/m^2^ [stage 0/B0]; diabetes 24.9% vs. 16.7% vs. 0%; hypertension 61.3% vs. 36.2% vs. 0%; and hyperlipidemia 31.6% vs. 23.7% vs. 7.7%).

**Table 1.** Demographic and clinical features of participants across heart failure stages.

|  | **Stage 0/B0** | **Stage A** | **Stage B1** |
| --- | --- | --- | --- |
| **n** | 558 | 527 | 253 |
| **Age, yrs (mean (SD))** | 41.2 (14.0) | 51.4 (15.0) | 58.1 (14.2) |
| **Sex (n (% female))** | 296 (53.0) | 284 (53.9) | 137 (54.2) |
| **Self-reported race (n (%))** |  |  |  |
| **Asian** | 102 (18.3) | 36 ( 6.8) | 19 (7.5) |
| **Black/African-American** | 54 (9.7) | 124 (23.5) | 76 (30.0) |
| **Other** | 64 (11.5) | 52 (9.9) | 14 (5.5) |
| **White** | 338 (60.6) | 315 (59.8) | 144 (56.9) |
| **Body mass index, kg/m<sup>2</sup> (mean (SD))** | 24.2 (2.9) | 31.1 (6.6) | 32.3 (6.8) |
| **History of diabetes (n (%))** | 0 (0.0) | 88 (16.7) | 63 (24.9) |
| **History of hypertension (n (%))** | 0 (0.0) | 191 (36.2) | 155 (61.3) |
| **History of hyperlipidemia (n (%))** | 43 (7.7) | 125 (23.7) | 80 (31.6) |
| **Epigenetic age acceleration, yrs (mean (SD))** | -0.24 (3.8) | -0.06 (4.2) | 0.47 (4.8) |

In the full analysis sample, we found an overall trend toward significant differences in EAA across HF stages (ANOVA p=0.07), with stage A (-0.1±4.2 years) showing intermediate values of EAA between stages 0/B0 (-0.2±3.8 years) and B1 (0.5±4.8 years) (**Figure 1A**). In sex-stratified models, the trend was similar in males (p=0.07), but there was no difference in EAA between HF stages in females (p=0.5) (**Figure 1B**). Among individual cardiac structural abnormality components comprising stage B HF, EAA was positively associated with left ventricular posterior wall diameter (β [95% CI]: 1.6 [0.03-3.1], p=0.046) (**Table 2**). Other echocardiographic parameters (including left ventricular mass index, relative wall thickness, left atrial volume index, E/e’ ratio) were not significantly associated with EAA. Finally, in analyses restricted to participants with stage A and B1 only (n=780), there was no association between EAA and stage B1 HF (β [95% CI]: 0.52 [-0.13-1.2], p=0.12). However, in sex-stratified analyses, males showed a marginal trend for EAA being associated with stage B1 vs. A HF, with higher EAA in stage B1 in males (n=359, β [95% CI]=0.97 [-0.006-1.9], p=0.051), while females showed no association (n=421, β [95% CI]=0.15 [-0.74-1.0], p=0.76).

**Table 2.** Associations of individual echocardiographic parameters with epigenetic age association.

| **Echo parameter** | **n** | **Regression estimate**<br>**(95% CI)** | **p-value** | **FDR-adjusted**<br>**p-value** |
| --- | --- | --- | --- | --- |
| **Average LV posterior wall diameter (cm)** | 1338 | 1.6 (0.027,3.1) | 0.046 | 0.36 |
| **LA volume index** | 1338 | 0.028 (-0.0031,0.058) | 0.078 | 0.36 |
| **LV relative wall thickness** | 1338 | 2 (-0.31,4.3) | 0.089 | 0.36 |
| **Average LV stroke volume (ml)** | 1208 | 0.0079 (-0.007,0.023) | 0.3 | 0.8 |
| **Average RV fractional area change (%)** | 1165 | 0.017 (-0.026,0.06) | 0.45 | 0.8 |
| **LV mass index** | 1338 | 0.005 (-0.0087,0.019) | 0.48 | 0.8 |
| **Average LV ejection fraction (%)** | 1208 | 0.018 (-0.043,0.08) | 0.55 | 0.8 |
| **RA area indexed** | 1236 | 0.038 (-0.099,0.17) | 0.59 | 0.8 |
| **LV end-systolic volume index** | 1208 | -0.0087 (-0.044,0.027) | 0.63 | 0.8 |
| **Average RV systolic area (cm<sup>2</sup>)** | 1165 | 0.022 (-0.079,0.12) | 0.67 | 0.8 |
| **LV end-diastolic volume index** | 1338 | -0.0018 (-0.019,0.016) | 0.84 | 0.9 |
| **RV area indexed** | 1165 | 0.0086 (-0.12,0.14) | 0.9 | 0.9 |

Regression estimates are derived from linear regression models adjusted for sex.
LV: left ventricular; LA: left atrial; RV: right ventricular; RA: right atrial

**Figure 1.**
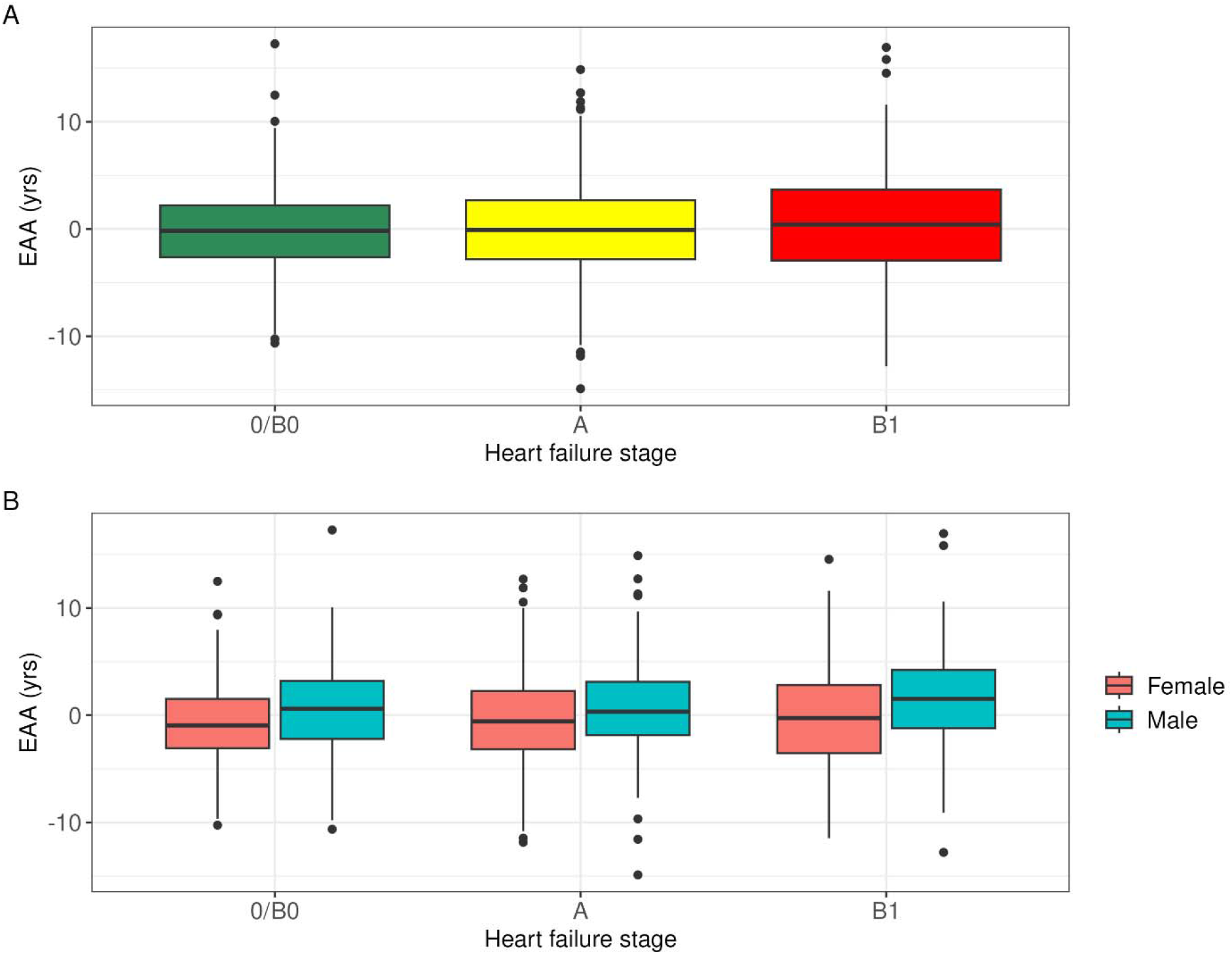
Distributions of epigenetic age acceleration (EAA) across heart failure stages for A) all included participants and B) participants stratified by sex.

## Discussion

Leveraging a deeply phenotyped cohort, we find a suggestion that in addition to previously-reported associations between EAA and symptomatic HF, EAA may also be associated with early stage HF. This suggests that biologic age acceleration captured by methylation may be higher in the presence of structural cardiac abnormalities (stage B1) as compared with risk factors alone (stage A). While EAA was associated with higher left ventricular posterior wall thickness, it was not associated with other structural changes, thus the mechanisms underlying the observed patterns may reflect broader cardiovascular aging processes, potentially involving multiple pathways beyond simple structural changes. Our results also suggest the presence of sex-specific effects, with males only demonstrating higher EAA in stage B1 as compared with stage A HF. While we were unable to account for menopausal status, given the mean age of our cohort these results could reflect protective effects of estrogens or, alternatively, less severe structural cardiac abnormalities in the setting of HF risk factors in females. Limitations of our study include the cross-sectional design, which precludes causal inferences, and potential residual confounding. Regardless, our study provides exploratory findings for a relationship between epigenetic age acceleration and early heart failure stages, especially in males. These findings support the potential role of epigenetic aging as a biomarker for early cardiovascular risk stratification. Future larger and longitudinal studies are needed to validate our findings and elucidate the temporal relationship between EAA and HF progression and to better understand the biological mechanisms underlying these associations.

## Acknowledgments

The authors wish to thank Project Baseline Health Study participants and study sites.

